# Distinct metastatic organotropism shapes prognosis in lung adenocarcinoma with brain metastasis

**DOI:** 10.1101/2025.02.25.25322889

**Authors:** Sama I. Sayin, Ella A. Eklund, Kevin X. Ali, Jozefina J. Dzanan, Moe Xylander, Martin Dankis, Per Lindahl, Volkan I. Sayin, Andreas Hallqvist, Clotilde Wiel

**Affiliations:** Department of Oncology, Sahlgrenska University Hospital, Gothenburg, Sweden; Sahlgrenska Center for Cancer Research, Department of Surgery, Institute of Clinical Sciences, University of Gothenburg, Gothenburg, Sweden; Wallenberg Centre for Molecular and Translational Medicine, University of Gothenburg, Gothenburg, Sweden; Department of Molecular and Clinical Medicine, Institute of Medicine, University of Gothenburg, Gothenburg, Sweden; Department of Biochemistry, Institute of Biomedicine, University of Gothenburg, Gothenburg, Sweden; Department of Oncology, Institute of Clinical Sciences, University of Gothenburg, Gothenburg, Sweden

**Keywords:** lung cancer, organotropism, brain metastasis, stage IV, LUAD

## Abstract

**Background:** Metastatic organotropism in lung cancer significantly influences prognosis, yet current treatment and clinical management guidelines are largely generalized for metastatic disease, regardless of organ site involvement. Notably, up to 30% of non-small cell lung cancer (NSCLC) patients present with brain metastases (BM) at diagnosis, underscoring the need for a more nuanced understanding of metastatic patterns. However, real-world clinical data on metastatic organotropism in well-characterized patient cohorts remain surprisingly scarce. Here, we evaluate patterns of metastasis, clinical characteristics and survival outcomes in patients with lung adenocarcinoma (LUAD), the major histological NSCLC subtype.

**Methods:** We performed a multi-center retrospective study including 913 stage IV LUAD patients, diagnosed and molecularly assessed in western Sweden between 2016 and 2021. Our primary study outcome was the distribution of specific metastatic sites and its impact on Overall Survival (OS).

**Results:** Out of 913 stage IV LUAD patients, 23.4% had BM. These patients exhibited markedly different metastatic patterns compared to those without BM, and median survival was significantly shorter (6 months) than those without BM (7.8 months) (*p =* 0.021). In addition, more than one metastatic tumor in the brain coincided with worse OS, compared to those with no, or with only one metastatic tumor in the brain. Importantly, OS was also influenced by metastasis in specific extracranial organs, like the pleura and lungs.

**Conclusions:** Our study highlights the distinct metastatic patterns and survival outcomes associated with BM in stage IV LUAD. These findings emphasize the need for site-specific approaches in managing metastatic disease due to the impact of BM on survival.

## Introduction

Lung cancer is the most prevalent and lethal cancer worldwide, and majority of patients are diagnosed with advanced metastatic disease (1). Non-small cell lung cancer (NSCLC) accounts for 85% of all cases, and among them lung adenocarcinoma (LUAD) is the most abundant histological subtype (1). While recent advances in treatment strategies like targeted- and immunotherapy have greatly improved outcomes for patients with early stage and locally advanced disease, no curative treatments exist till date for metastatic disease, which remains the leading cause of mortality in these patients (2). Identification of novel prognostic factors to further guide clinical management of metastatic disease is therefore crucial and urgent.

It is now well-established that metastatic dissemination of primary tumors throughout the body is not random, and solid tumors metastasize preferentially to certain organs, a process termed metastatic organotropism. While this phenomenon has been studied extensively in animal models (3, 4), few studies until recently have reported metastatic organotropism patterns of lung cancer in the clinic setting. Emerging evidence now suggests that metastatic organotropism in lung cancer patients is strongly related to previously well-established prognostic factors such as age (5, 6), oncogenic driver mutations (7), histological subtypes (8), as well as response to treatment (5, 7, 9).

Importantly, current treatment and clinical management recommendations apply broadly for metastatic disease independent of organ site involvement (10). While this can be followed in practice for metastasis to other organs, metastatic involvement of the brain requires specialized treatment and management strategies in clinical reality, owing to the sensitivity of the anatomical location and the highly selective permeability of the blood-brain barrier to systemic treatment agents (11). Nevertheless, BM is reported at diagnosis in 25-29% of NSCLC patients with metastatic disease and up to 50% will develop BM during the disease course (12).

Importantly, patients with BM have significantly worse prognosis than those with only extracranial metastatic disease (11, 13-15). In addition, BM patients with stable intracranial metastatic disease who have progressive extracranial disease have worse prognosis than those with stable extracranial disease(16). Metastatic involvement of certain extracranial organs but not others have been shown to affect response to immunotherapy among BM patients (17, 18). While preclinical studies have provided possible explanations for these differences through demonstrating unique biological phenotypes of BM compared to primary tumors and their metastases in other organ sites (7, 19), clinical data to aid further stratification within BM patient group to guide clinical management based on extracranial metastatic disease patterns are lacking.

Real-world clinical data on metastatic organotropism in well-characterized patient cohorts are surprisingly scarse, and studies of organotropism in relation to BM and its effect on clinical outcomes are lacking. Here, we report in detail metastatic organotropism in relation to BM and related clinical outcomes in all patients with stage IV/metastatic lung adenocarcinoma (LUAD) in the West Sweden cohort (9, 20). We present a comprehensive report on patterns of metastasis, clinical characteristics and survival outcomes in western Sweden by combining data from the Swedish Lung Cancer Registry (SLCR) with 95% coverage, manual health chart data curation and histopathological analyses.

## Materials and Methods

By combining data about metastatic sites recorded in the SLCR and through data curation from health charts to identify sites of metastasis unreported/not included in original report. The Swedish healthcare system is primarily government-funded and provides universal access to all citizens. Therefore, patients have equal access to diagnostic examinations and treatments.

### Patient Population

We conducted a multi-center retrospective study including all consecutive NSCLC patients diagnosed with Stage IV LUAD and having molecular assessment performed between 2016– 2021 in western Sweden (*n* = 913). Patient demographics (including age, gender, Eastern Cooperative Oncology Group (ECOG) performance status and smoking history), cancer stage, sites of metastasis, pathological details (histology, mutation status) and outcome data were retrospectively collected from patient charts and the Swedish Lung Cancer Registry. Approval from the Swedish Ethical Review Authority (Dnr 2019-04771) was obtained prior to study commencement.

### Study Objectives

The primary outcome of this study was presence of metastasis in given organ sites at diagnosis and overall survival (OS), defined as the interval between the date of diagnostic sample collection and the date of death from any cause. Patients alive or lost to follow-up were censored at the cut-off date or last contact. Median follow-up time was 35 months (95% CI 31.1–38.9) and was estimated using the reverse Kaplan–Meier method. One patient died before final diagnosis and is thus excluded from the OS analysis. We compared OS stratified on *metastatic organ involvement* for the entire cohort. BM diagnosed within 8 weeks from date of diagnostic sample collection was considered as diagnosed at baseline. Data cut-off date was 2024-09-17.

### Statistical Analysis

Clinical characteristics were summarized using descriptive statistics and evaluated with univariate analysis in table form. Distribution of metastatic sites was assessed with Pearson Correlation. Survival was estimated using the Kaplan–Meier method. Log-rank test was used to assess significant differences in OS between groups. Multivariable Cox regression analyses were conducted to compensate for potential confounders. Statistical significance was set at *p* < 0.05, and no adjustments were made for multiple comparisons. Data analysis was conducted using IBM SPSS Statistics version 27 and R version 3.4.

## Results

### Patient characteristics

All consecutively diagnosed patients with LUAD in West Sweden between 2016-2021 with molecular assessment were included in this study. Total 913 patients with stage IV LUAD were included in the study (Figure 1) and clinical characteristics are summarized in Table 1.

**Table 1.** Characteristics of the total cohort as well as stratified by presence or absence of brain metastasis at diagnosis.

| Patient characteristics | No Brain Metastasis | Brain Metastasis | Total | p-value |
| --- | --- | --- | --- | --- |
| **All subjects** | n=699 | n=214 | n=913 |  |
| Median age at diagnosis (min, max) | 72 (26, 94) | 70 (34, 87) | 72 (26, 94) | 0.640 |
| Median survival in months (95% CI) | 7.8 (6.5-9.0) | 6.0 (4.5-7.5) | 7.1 (6.2-8.0) | **0.021** |
| **Sex (%)** |  |  |  | **0.028** |
| Male | 324 (46.6) | 81 (37.9) | 405 (44.4) |  |
| Female | 375 (53.6) | 133 (62.1) | 508 (55.6) |  |
| **Alive at follow-up (%)** |  |  |  | 0.358 |
| Alive | 67 (9.6) | 17 (7.9) | 83 (9.1) |  |
| Deceased | 632 (90.4) | 197 (92.1) | 830 (90.9) |  |
| **Smoking history (%)** |  |  |  | 0.158 |
| Current smoker | 204 (29.2) | 77 (36.0) | 281 (30.8) |  |
| Former smoker | 342 (48.9) | 96 (44.9) | 438 (48.0) |  |
| Never smoker | 151 (21.6) | 40 (18.7) | 191 (20.9) |  |
| Missing | 2 (0.3) | 1 (0.5) | 3 (0.3) |  |
| **Performance status (%)** |  |  |  | 0.316 |
| Grade 0 | 74 (10.6) | 27 (12.6) | 101 (11.1) |  |
| Grade 1 | 258 (36.9) | 83 (38.8) | 341 (37.3) |  |
| Grade 2 | 184 (26.3) | 61 (28.5) | 245 (26.8) |  |
| Grade 3 | 120 (17.2) | 24 (11.2) | 144 (15.8) |  |
| Grade 4 | 26 (3.7) | 8 (3.7) | 34 (3.7) |  |
| Missing | 37 (5.3) | 11 (5.1) | 48 (5.3) |  |
| **Mutation status (%)** |  |  |  |  |
| KRAS | 265 (37.9) | 72 (33.6) | 337 (36.9) | 0.258 |
| KRAS-G12C | 106 (15.2) | 34 (15.9) | 140 (15.3) | 0.298 |
| EGFR | 97 (13.9) | 49 (22.9) | 146 (16) | **0.002** |
| ALK | 25 (3.6) | 9 (4.2) | 34 (3.7) | 0.671 |
| ROS1 | 22 (3.1) | 1 (0.5) | 23 (2.5) | **0.029** |
| BRAF | 39 (5.6) | 5 (2.3) | 44 (4.8) | 0.053 |
| **PDL1-grade (%)** |  |  |  | 0.500 |
| 0% | 356 (50.6) | 117 (54.7) | 471 (51.6) |  |
| ≥1% | 116 (16.5) | 26 (12.1) | 140 (15.3) |  |
| ≥20% | 65 (9.2) | 20 (9.3) | 85 (9.3) |  |
| ≥50% | 167 (23.7) | 51 (23.8) | 217 (23.8) |  |

**Figure 1.**
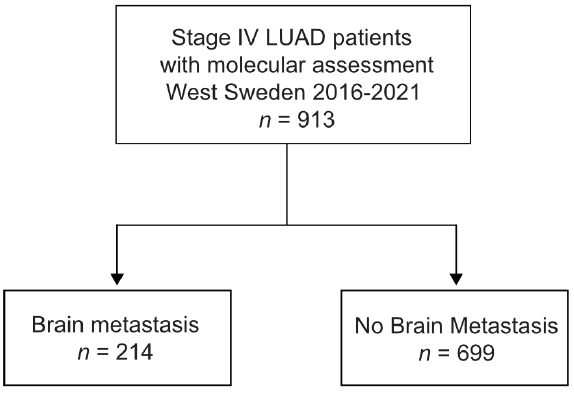
Patient Selection. Flow chart showing patient selection for the study.

Of all patients with metastatic disease, those with BM had significantly lower median survival (6 months) than those without BM (7.8 months) (*p =* 0.021). There were significantly higher proportion of females in the BM group (62.1%) than No BM group (53.6%) (*p =* 0.028). KRAS was the most frequently mutated gene in primary tumors of both groups, while significantly higher proportion of BM patients had mutations in EGFR (22.9% vs 13.9%; *p =* 0.002).

### Metastatic organotropism in Stage IV lung adenocarcinoma

First, we mapped the distribution of metastatic sites for each patient with stage IV disease. We identified subgroups by specific organs involved and mapped individual pattern for each patient (Figure 2 and Supplementary Figure 1). Most patients had metastasis to the bone (38.6%, *n =* 352), followed by the lung (27.8%, *n =* 254), pleura (24.8%, *n =* 226) and brain (23.4%, *n =* 214) (Figure 3A and Supplementary Figure 2A). Liver was the site with the least number of patients with metastatic involvement (13.8%, *n =* 94) followed by adrenal gland (16.9%, *n =* 123). Importantly, up to 54% of patients with pleura metastasis had no other metastatic organ involvement, followed by lung (44.1%) and brain (39.9%) as sites with higher levels of single organ metastasis. In contrast, only 22.7% of all patients with liver- and 15.7% with adrenal metastasis did not involve any other organs (Figure 3B).

**Figure 2.**
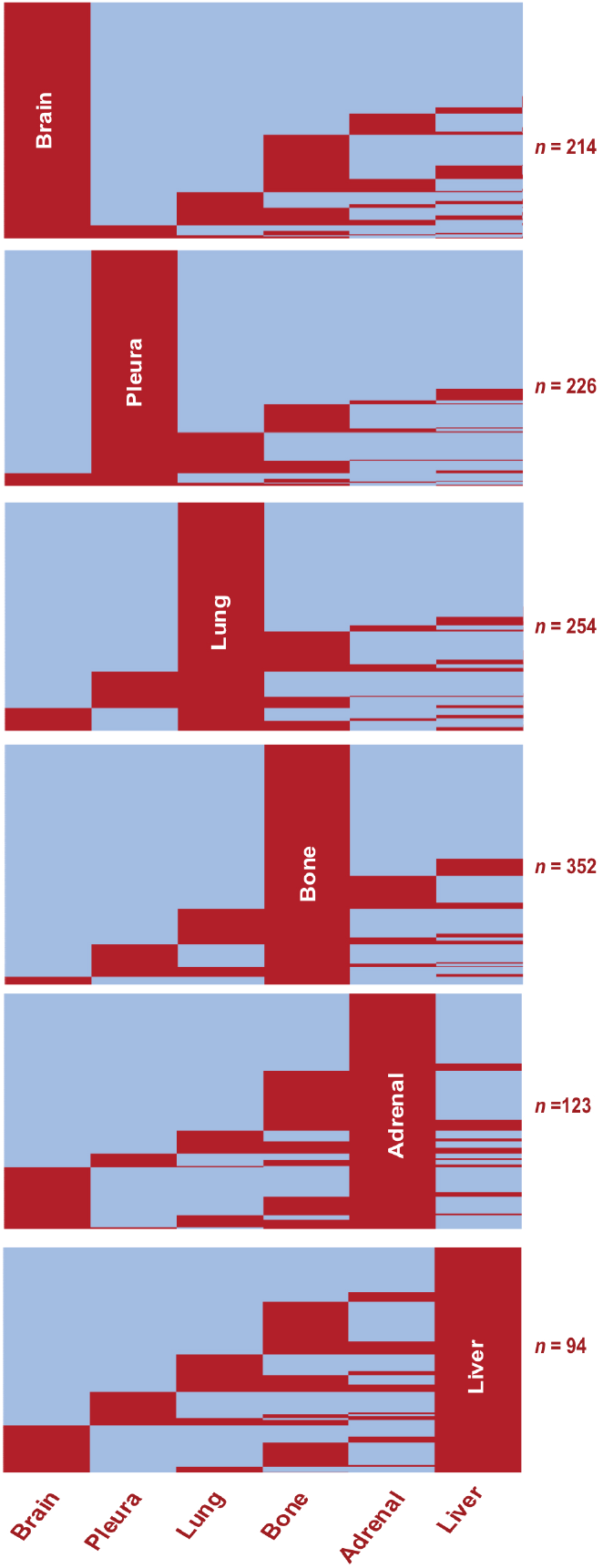
Metastatic Organotropism in Stage IV LUAD. Heatmaps showing presence (red) or absence (blue) of metastasis at given organ sites in the study population (*n* = 913). Rows represent individual patients. Subgroups show pattern of metastasis among all patients with the given organ site involvement.

**Figure 3.**
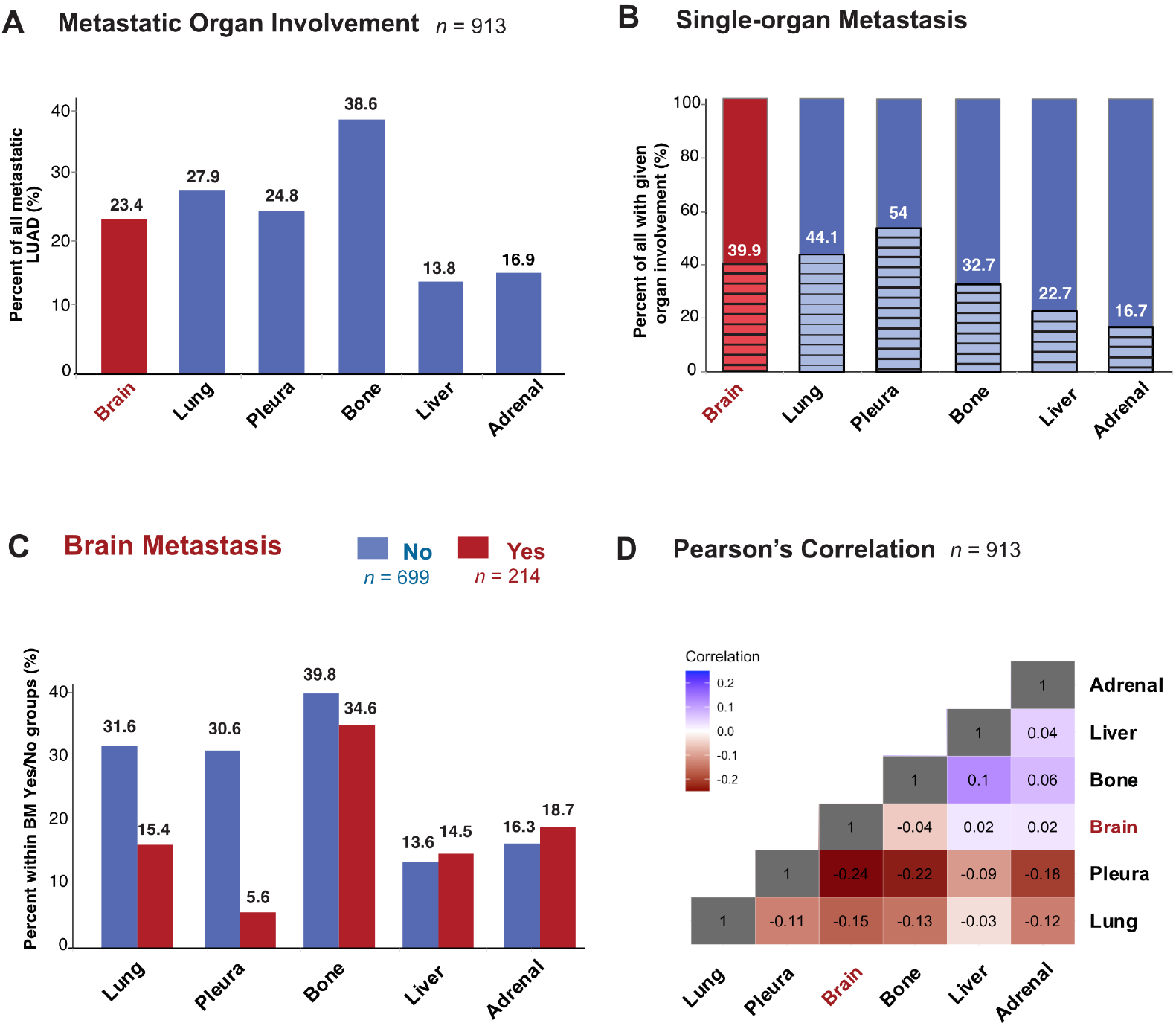
Metastatic Organotropism in relation to BM in stage IV LUAD. **(A)** Percentage of all patients in the study population with metastatic involvement of given organ site at diagnosis. **(B)** Percentage of all patients with given organ site involvement with single-organ metastasis **(C)** Percentage with metastatic involvement of given organ site at diagnosis in subgroups with BM (red) and without BM (blue). **(D)** Heatmap showing Pearson’s correlation values between pairs of organ sites of metastasis.

### Metastatic organotropism of the lung and pleura, but not bone, liver and adrenal is altered in BM patients

Next, to study organotropism of LUAD in relation to the brain, we first sub-grouped patients into those with (*n =* 214) or without (*n =* 699) BM (Figure 3C). Bone was the organ of metastasis for most patients with (34.6%) or without BM (39.8%). The percentage of patients with metastasis to the liver and adrenal were similar regardless of brain involvement. In contrast to the bone, adrenal and liver, where the proportions were similar between BM or no BM groups, there were large differences in metastasis to the pleura and lung depending on brain involvement status. The most dramatic difference was seen in patients with BM, who unlike those without BM, had only 5.6% metastasis to the pleura, while those without BM 30.6% had metastasis to the pleura. Similarly, in the lung, while metastatic involvement in no BM group was 31.6%, only 15.4% of stage IV LUAD with metastases in the brain also had metastases in the lung. Pearson’s correlation coefficient was accordingly lowest (−0.24) between the pleura and brain, while liver and bone had the highest correlation (0.1) (Figure 3D and Supplementary Figure 2B).

### Metastasis to the brain affects survival outcomes in LUAD

We found that among all patients with metastasized LUAD, presence of metastasis in the brain correlated significantly (*p =* 0.019) with worse OS (6.0 months; 95% CI 4.5-7.5) compared with patients with metastatic disease without brain involvement (7.8 months; 95% CI 6.5-9.1) (Figure 4A). Interestingly, having more than one metastatic tumor in the brain corresponded with significantly worse OS (4.7 months; 95% CI 3.0-6.4) compared to those without (7.8 months; 95% CI 6.5-9.1) or with only one metastatic lesion in the brain (8.1 months; 95% CI 5.4-10.9) (Figure 4B). Multivariate analysis showed BM as the second most significant variable affecting OS, with less effect than ECOG but similar effect to smoking and greater effect than age at diagnosis on survival outcomes (Figure 4C).

**Figure 4.**
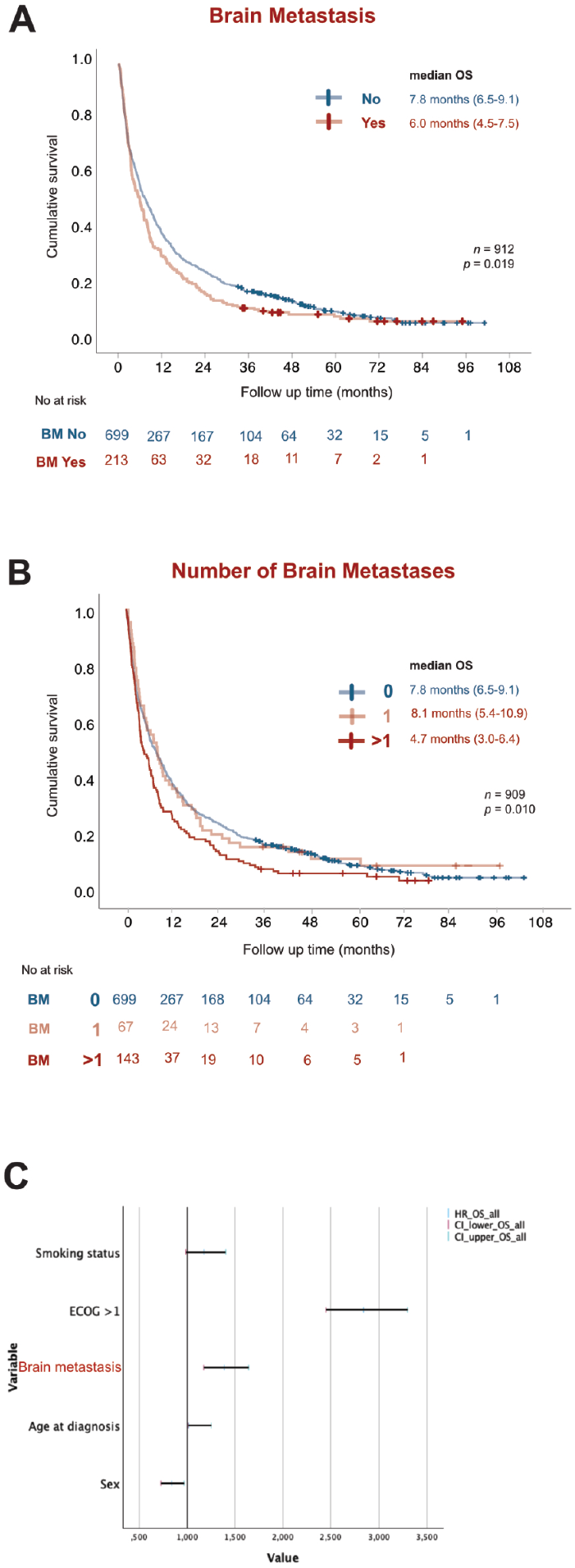
BM worsens prognosis in stage IV LUAD. Kaplan-Meier estimates comparing **(A)** overall survival (OS) stratified by presence (red) or absence (blue) of BM. **(B)** OS stratified by total number of BM present at diagnosis as no brain metastasis (blue), 1 BM tumor (light red) and more than 1 BM tumor (dark red). **(C)** Forest plot of multivariate COX regression analysis for overall survival in the study population. OS: overall survival; NR Not reached; HR: Hazard Ratio; CI: Confidence of interval.

### Metastatic organotropism in relation to BM affects survival outcomes in LUAD

Next, we analyzed how metastatic involvement of each organ site in relation to BM affected survival outcomes. Importantly, metastatic involvement of each organ in relation to the brain had significant effects on OS (Figure 5 and Table 2). For all organs, BM patients had worse OS than those with no BM independent of other organ involvement. Among BM patients, involvement of pleura, lung or liver worsened prognosis independent of other organ involvement, while metastatic involvement of the bone or adrenal gland did not affect survival further among BM patients (Figure 5 and Table 2).

**Fig. 5.**
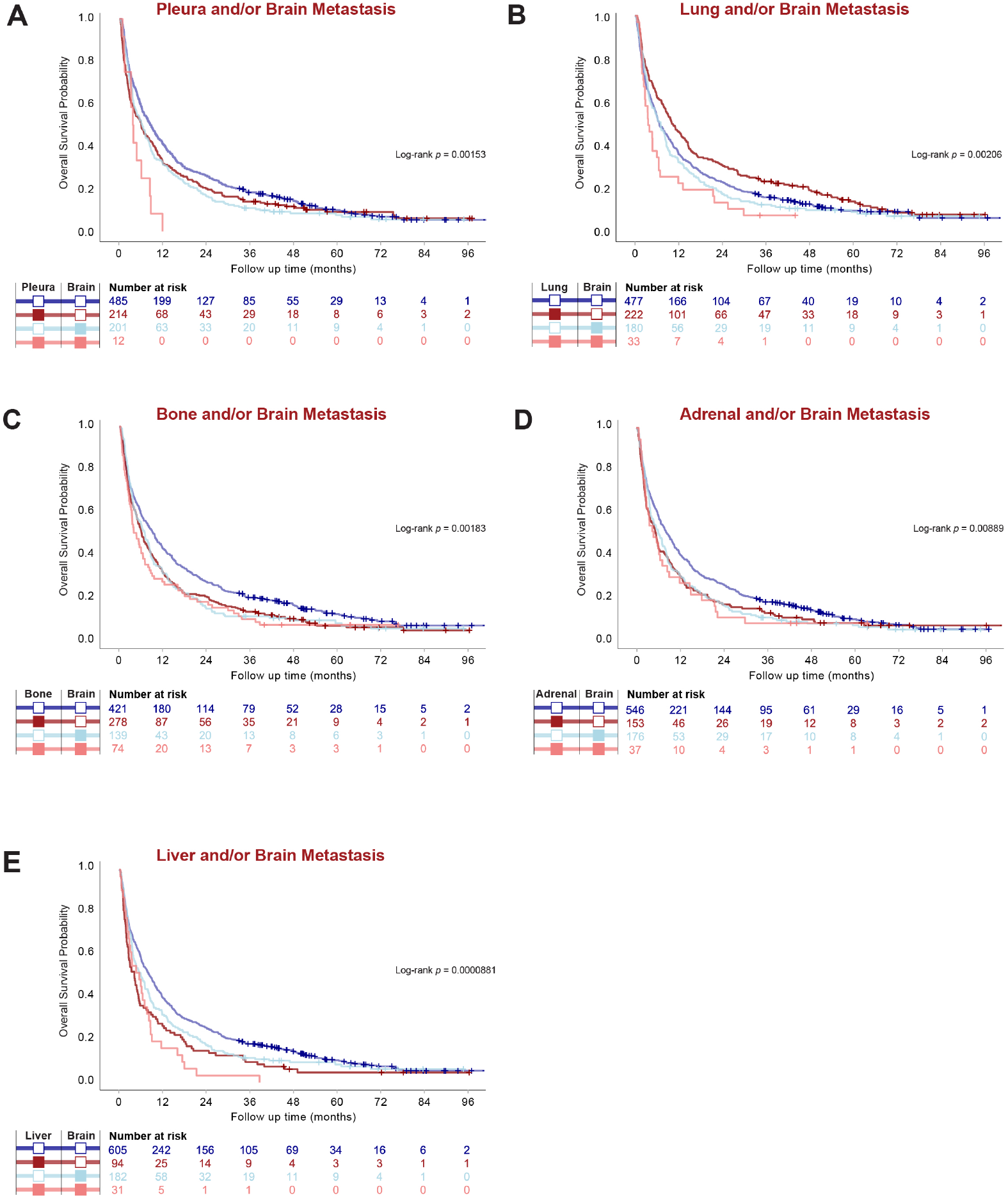
Metastatic Organotropism in relation to BM affects survival outcomes in stage IV LUAD (A-E) Kaplan-Meier estimates comparing overall survival (OS) by given organ-site involvement in relation to BM.

**Table 2.**
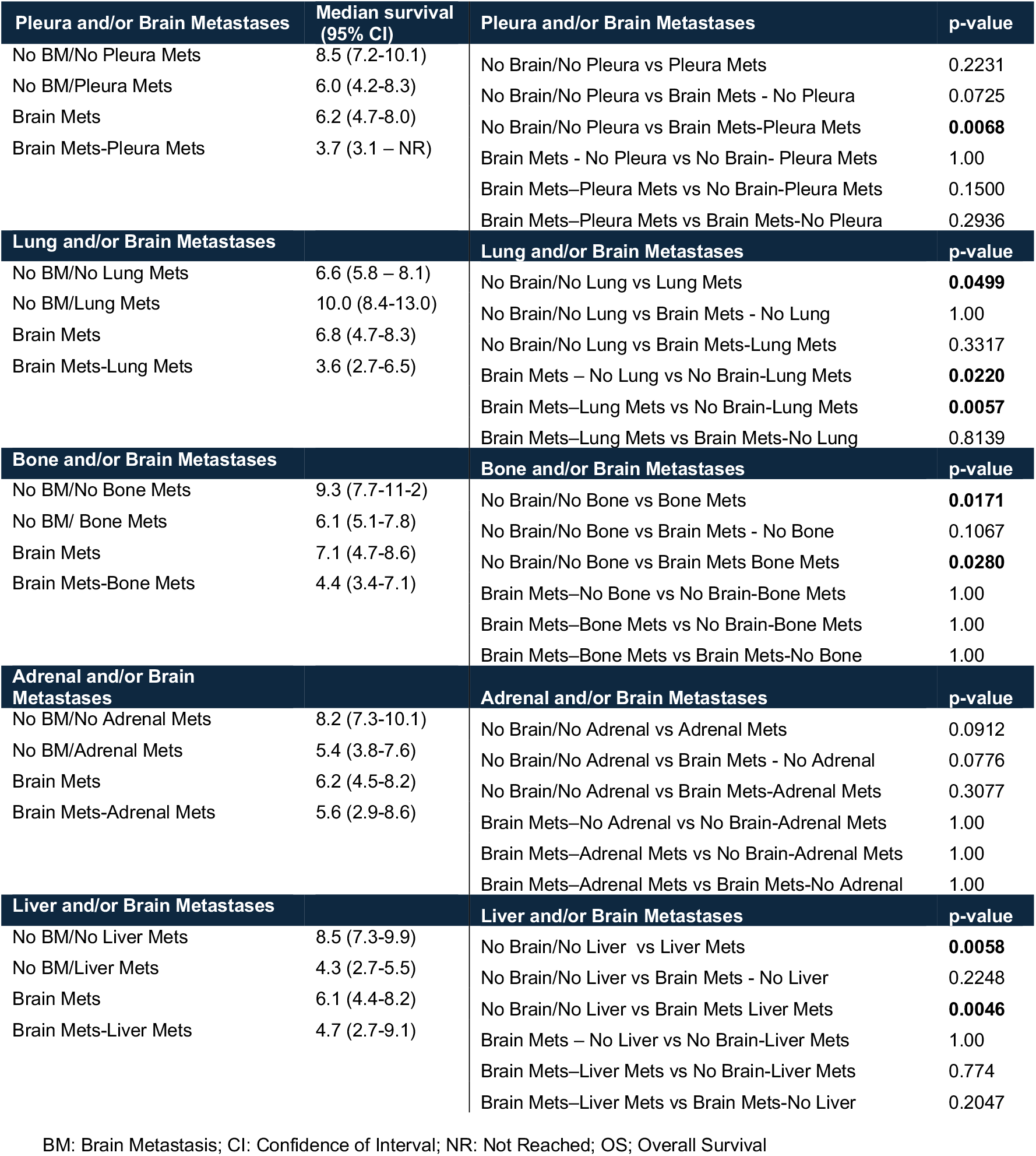
Median survival and individual p-values for pairwise comparison of OS presented in Figure 5.

### BM patients with pleura or lung metastasis have worse prognosis

BM patients with pleura metastasis had drastically worse OS (3.7 months; 95% CI 3.1-NR) than those without pleura involvement (6.2 months; 95% CI 4.7-8.0), while pleura metastasis did not affect survival in the absence of BM (*p =* 0.2231). Interestingly, BM patients with metastasis to the lungs had numerically worse OS (3.6 months; 95% CI 2.7-6.5) than BM patients (6.8 months; 95% CI 4.7-8.3), although metastasis to the lungs in the absence of BM even improved OS (10.0 months; 95% CI 8.4-13.0) (*p =* 0.0499) (Figure 5 and Table 2).

## Discussion

This multicenter retrospective study provides novel insights into metastatic organotropism in LUAD with a specific focus on BM and their impact on clinical outcomes. Our findings reveal several critical and previously unreported aspects of metastatic patterns and survival in patients with stage IV LUAD, particularly highlighting the distinct organotropism of metastasis in relation to the brain. Notably, we found that patients with BM exhibit significantly altered metastatic patterns compared to those without BM, characterized by a markedly lower prevalence of pleura and lung metastases. Furthermore, our analysis demonstrated that BM is associated with worse OS, and that survival outcomes are further modulated by metastatic involvement of specific extracranial organs, such as the pleura and lungs. These findings provide compelling evidence of the unique biology underlying brain metastases in LUAD and emphasize the critical need for organ site-specific approaches in the management of metastatic disease.

We and others have previously reported higher frequency of BM in females with NSCLC, consistent with the findings in the present study (21, 22). Our results also corroborate previous reports showing that BM is associated with lower median survival (15) and a higher frequency of EGFR mutations (23-26), further emphasizing the clinical and molecular uniqueness of BM in LUAD. To our knowledge, the negative correlation between metastasis to the brain and the pleura or lungs observed in this study has not been reported previously. This unique finding suggests that the presence of BM may actively influence metastatic patterns, potentially limiting involvement of certain organ systems such as the pleura.

The frequency distribution of metastatic sites in our cohort aligns with previous studies by Tamura et al. and Lengel et al., where the highest rates of metastases were observed in the bone, lung, brain, adrenal gland, and liver in descending order (7, 27). Similar to these studies, our data also show that the liver and adrenal gland had the lowest frequency of metastatic involvement and were also the least likely sites of single-organ metastases. These findings could suggest that metastases to these organs may occur later in the metastatic cascade, supporting the hypothesis that they represent more advanced disease stages in LUAD.

In contrast to Tamura et al. (27), who reported that liver and adrenal metastases were associated with poor survival in NSCLC but found no significant survival differences related to lung and pleura involvement when studied independent of BM, our study highlights the critical prognostic impact of pleura and lung metastases in the presence of BM. Specifically, we found that while pleura metastases alone did not affect survival, their coexistence with BM drastically worsened prognosis, further underscoring the importance of organ-specific interactions in metastatic disease.

Rihimäki et al. (28) previously reported metastatic site involvement of the nervous system and respiratory system using data from the Swedish Cancer Registry and the Swedish National Cause of Death Registry. However, our analysis by combining more detailed-level data from the Swedish Lung Cancer Registry and manual curation of health records maps these sites of metastasis with higher resolution and accuracy. This distinction reveals a survival advantage associated with lung metastases in patients without BM, as also reported by Li et al. (5), but highlights a negative prognostic impact of lung metastases in the presence of BM.

Finally, we acknowledge several clinical and methodological limitations inherent to this and other studies of metastatic organotropism. First, we have only studied metastatic site involvement at diagnosis. Metastatic involvement is generally underreported, especially for sites that become involved later during the disease course or during palliative treatment, as metastatic sites are not routinely mapped without clinical indication. Additionally, autopsy studies frequently identify previously undetected metastatic sites, leading to discrepancies in reported frequencies compared to those based on diagnosis alone (29). These factors highlight the need for comprehensive and standardized approaches to map metastatic organotropism across the disease trajectory.

## Conclusion

This study demonstrates that metastatic organotropism differs significantly among LUAD patients depending on the presence of brain metastases. Notably, metastases to the pleura and lung are rare but drastically worsen prognosis in BM patients, suggesting unique organ-specific interactions that influence survival outcomes. These findings provide novel insights into the biology of metastatic spread in LUAD and underscore the critical need for organ-specific treatment strategies, particularly in the management of brain metastases. Further research is warranted to elucidate the underlying mechanisms driving these patterns and to optimize clinical management for patients with advanced disease.

## Abbreviations

BM: Brain Metastasis
ECOG: Eastern Cooperative Oncology Group
HR: Hazard Ratio
LUAD: Lung Adenocarcinoma
NSCLC: Non-Small Cell Lung Cancer
OS: Overall Survival
PS: Performance Status

## Declarations

## Acknowledgments

We thank members of the Swedish Lung Cancer Registry and the continuous reporting by Swedish healthcare employees.

## Authors’ contributions

Conceptualization, E.A.E., S.I.S., A.H., V.I.S. and C.W; Data curation, S.I.S., E.A.E.; Formal analysis, E.A.E., S.I.S.; K.X.A., J.J.D., M.D., Funding acquisition, E.A.E., C.W., A.H. and V.I.S.; Methodology, S.I.S., E.A.E.; Supervision, S.I.S., A.H, V.I.S. and C.W; Visualization, S.I.S., K.X.A., M.X.; Writing—original draft, S.I.S., E.A.E. and C.W.; Writing—review & editing, S.I.S, E.A.E., K.X.A, J.J.D., M.X., M.D., P.L., V.I.S., A.H. and C.W, Project coordination, V.I.S. and C.W. All authors have read and agreed to the published version of the manuscript.

## Institutional Review Board Statement

Approval from the Swedish Ethical Review Authority (Dnr 2019-04771 and 2021-04987) was obtained prior to the commencement of the study. No informed consent was required due to all data presented in a de-identified form according to the Swedish Ethical Review Authority.

## Consent for publication

Not applicable. Patient consent statements were not required due to the retrospective nature of this study. No informed consent was required due to all data presented in a de-identified form according to the Swedish Ethical Review Authority.

## Data Availability Statement

The datasets used and/or analysed during the current study available from the corresponding author on reasonable request.

---

**Supplementary Figure 1.**
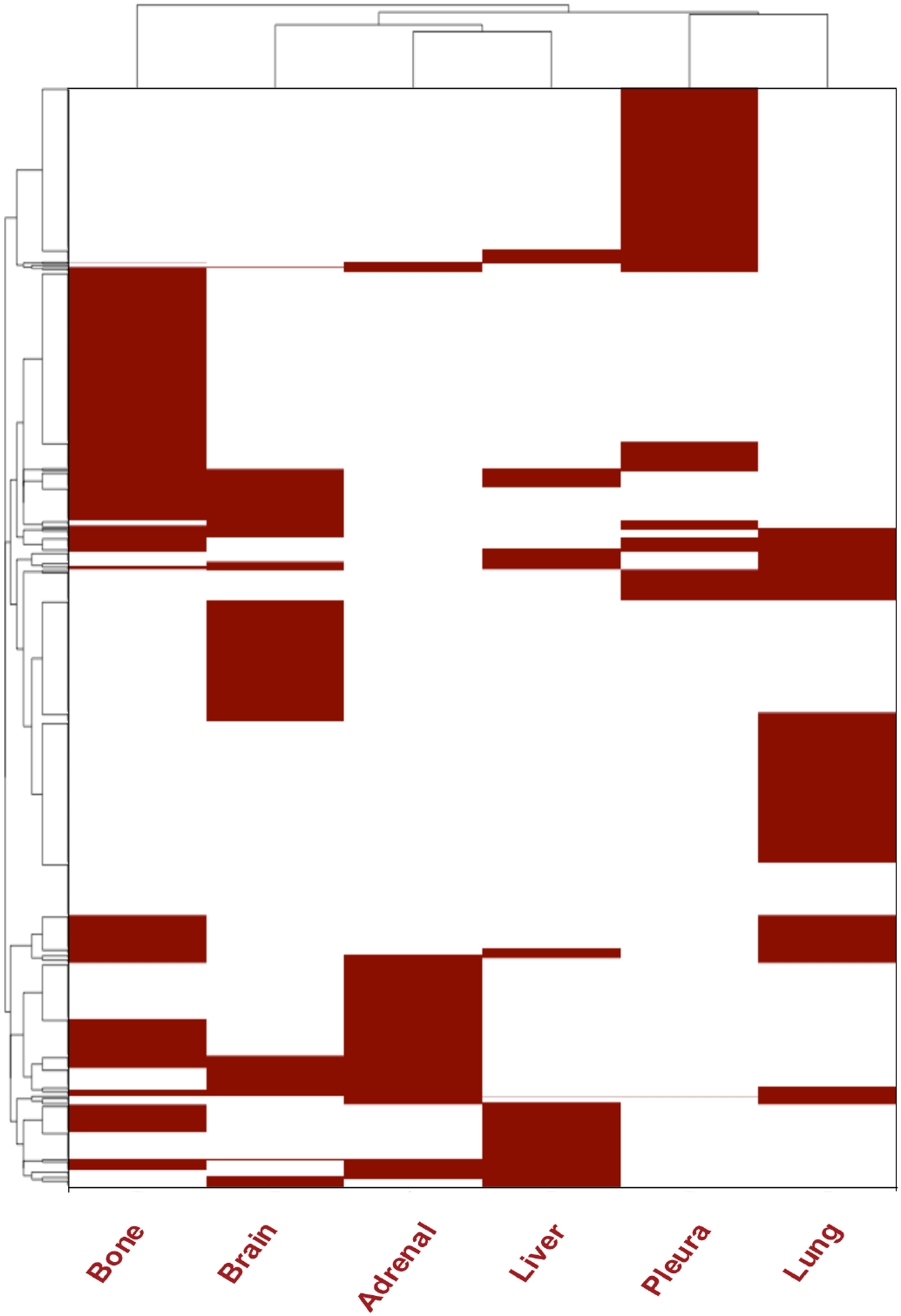
Distribution of most frequent sites of metastasis in stage IV LUAD. Heatmap showing unsupervised hierarchical clustering of sites of metastasis in the entire study population (*n* = 913). Rows represent individual patients.

**Supplementary Figure 2.**
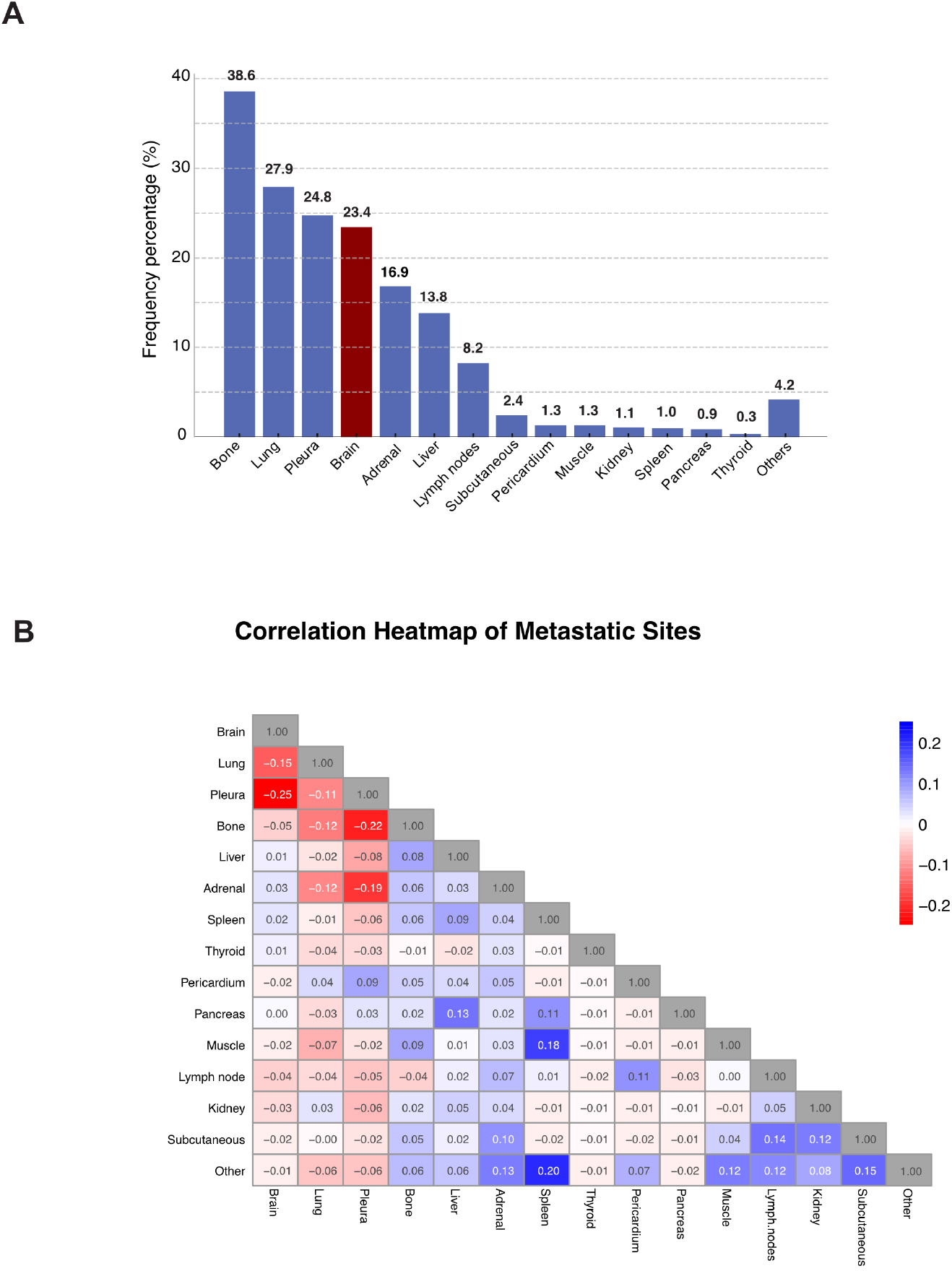
(**A)** Distribution of all sites of metastasis in Stage IV LUAD, including less frequent sites **(B)** Pearson chart showing correlation coefficient between all organ sites of metastasis.

